# The seroprevalence of SARS-CoV-2-specific antibodies in Australian children: a cross sectional study

**DOI:** 10.1101/2024.03.03.24303672

**Authors:** Archana Koirala, Jocelynne McRae, Philip N Britton, Marnie Downes, Shayal A Prasad, Suellen Nicholson, Noni E Winkler, Matthew V N O’Sullivan, Fatima Gondalwala, Cecile Castellano, Emma Carey, Alexandra Hendry, Nigel Crawford, Ushma Wadia, Peter Richmond, Helen S Marshall, Julia E Clark, Joshua R Francis, Jeremy Carr, Adam Bartlett, Brendan McMullan, Justin Skowno, Donald Hannah, Andrew Davidson, Britta S von Ungern-Sternberg, Paul Lee-Archer, Laura L Burgoyne, Edith B Waugh, John B Carlin, Zin Naing, Nicole Kerly, Alissa McMinn, Guillian Hunter, Christine Heath, Natascha D’Angelo, Carolyn Finucane, Laura A Francis, Sonia Dougherty, William Rawlinson, Theo Karapanagiotidis, Natalie Cain, Rianne Brizuela, Christopher C Blyth, Nicholas Wood, Kristine Macartney

## Abstract

**Background:** Following reduction of public health and social measures concurrent with SARS-CoV-2 Omicron emergence in late 2021 in Australia, COVID-19 case notification rates rose rapidly. As rates of direct viral testing and reporting dropped, true infection rates were most likely to be underestimated.

**Objective:** To better understand infection rates and immunity in this population, we aimed to estimate SARS-CoV-2 seroprevalence in Australians aged 0-19 years.

**Methods:** We conducted a national cross sectional serosurvey from June 1, 2022, to August 31, 2022, in children aged 0-19 years undergoing an anesthetic procedure at eight tertiary pediatric hospitals. Parents or guardians of children and adolescents under 18 years provided written consent and participants aged 18-19 years provided their own consent. Participant questionnaires were administered, and blood samples tested using the Roche Elecsys Anti-SARS-CoV-2 total spike and nucleocapsid antibody assays. S and N seroprevalence adjusted for geographic and socioeconomic imbalances in the participant sample compared to the Australian population was estimated using multilevel regression and poststratification within a Bayesian framework.

**Results:** Blood was collected from 2,046 participants (median age: 6.6 years). Adjusted seroprevalence of spike-antibody was 92.1 % (95% credible interval (CrI) 91.0-93.3%) and nucleocapsid-antibody was 67.0% (95% CrI 64.6-69.3). In unvaccinated children spike and nucleocapsid antibody seroprevalences were 84.2% (95% CrI 81.9-86.5) and 67.1% (95%CrI 64.0-69.8), respectively. Seroprevalence increased with age but was similar across geographic distribution and socioeconomic quintiles.

**Conclusion:** Most Australian children and adolescents aged 0-19 years, across all jurisdictions were infected with SARS-CoV-2 by August 2022, suggesting rapid and uniform spread across the population in a very short time period. High seropositivity in unvaccinated children informed COVID-19 vaccine recommendations in Australia.

**Funding:** Australian Government Department of Health and Aged Care.

## Introduction

Understanding Severe Acquired Respiratory Syndrome Coronavirus (SARS-CoV-2) population-level infection rates is important to order to inform infection related risk and contextualize severe outcome rates, such as hospitalization and death. Surveillance for SARS-CoV-2 based on case reporting and hospital notification data underestimates the true number of SARS-CoV-2 infections(1). Children in particular are subject to underreporting of infections due to higher rates of asymptomatic or mild infections(2–5) and difficulties in testing(6) but provide a unique population to understand spread, given lower vaccination rates in children compared to adults and infants < 2 years being born since the 1^st^ Omicron wave. Serosurveillance, which measures serum antibodies in individuals within a population, can give insights into the cumulative prevalence of infection and/or vaccine uptake over time across a population.

Methods for obtaining blood samples can include residual sera from diagnostic pathology laboratories, blood donors or antenatal collections. However, such approaches result in children being underrepresented due to the lack of routine blood testing, lack of adequate volumes of residual sera and/or reluctance to submit children to painful procedures such as venipuncture, without clear necessity. Obtaining child population representative blood sampling is difficult and costly, resulting in small non representative cohorts. Of 4,160 serosurveys logged on SeroTracker globally: only 323 serosurveys have examined children.(2) In 2020, a serosurveillance study conducted in New South Wales, the most populous state in Australia, using residual diagnostic sera for testing found that children 0-9 years were the only age group in which recruitment targets were unable to be met(6).

An Australian pediatric hospital sentinel surveillance program, the Paediatric Active Enhanced Disease Surveillance network (PAEDS), has been operating since 2007. The network consists of eight tertiary pediatric hospitals, across five states and one territory that assess emergency department presentations and hospitalizations of select key communicable diseases(1, 7, 8) and associated syndromes, including COVID-19,(9, 10) and Multisystem inflammatory Syndrome in children (or Pediatric inflammatory multisystem syndrome – temporally associated with SARS-CoV-2)(11, 12). Using this network, we previously conducted a SARS-CoV-2 serosurvey in children undergoing an elective anesthetic procedure within PAEDS hospitals, between November 2020 to March 2021. From a sample of 1685 children aged 0-19 years, the national seroprevalence of SARS-CoV-2 spike antibody (S-antibody) was estimated to be <0.6%(13), consistent with evidence of limited transmission of SARS-CoV-2 in Australia until June 2021(14). This survey(13) sampled children from all socioeconomic groups, rural and regional areas, and was broadly representative of the Australian population.

Following the emergence of the Omicron variant and easing of social and public health restrictions, very high SARS-CoV-2 case notifications occurred across all age groups in Australia from December 2021(15). Case notification data were not a reliable reflection of infection rates in the population, as tests were costly and required infected individuals or carers to self-notify their positive results.

Vaccines for SARS-CoV-2 became available for adults (including pregnant women) from 1 February 2021, in adolescents 12-15 years from 13 September 2021 and for children aged 5-11 years from 20 January 2022.(16) Vaccines used were mRNA based: Pfizer Cominarty (≥ 5 years) and Moderna Spikevax (≥ 6 years); the adenoviral-vectored vaccine: AstraZeneca Vaxzevria (adults ≥ 18 years), and the protein subunit vaccine, Novavax Nuxavoid (≥ 12 years). All available vaccines induced a S-antibody response, but no nucleocapsid antibody (N-antibody) response. As SARS-CoV-2 infection induced a S- and N-antibody response, presence of N-antibody was a marker of infection in those vaccinated. Presence of S-and N-antibody in infants could represent infection or maternal antibody transferred to the fetus during pregnancy. Sequential serosurveys in Australian adult blood donors reported S-antibody positivity rates of > 90% and N-antibody positivity rates that rose from 17% in February to 46% by June 2022(17), but no data were available in children. We aimed to estimate SARS-CoV-2 S-and N-antibody seropositivity to calculate a more accurate estimate population infection and hospitalization rates in Australian children and young adults aged 0-19 years after the emergence of the Omicron variant, vaccine rollout and opening of internal borders and easing of public health restrictions in Australia. We also aim to describe S and N-antibody levels in infected children who completed a primary 2-dose vaccination schedule versus unvaccinated children, and describe features of COVID-19 hospitalization in the serosurvey participants.

## Material and Methods

### Participants, study setting and recruitment

Individuals aged 0-19 years were recruited prior to undergoing an elective surgical procedure requiring general anesthesia at one of eight pediatric tertiary referral hospitals across six of eight Australian jurisdictions: Queensland Children’s Hospital, Brisbane, Queensland; Sydney Children’s Hospital Randwick and the Children’s Hospital at Westmead, Sydney, New South Wales; The Royal Children’s Hospital and Monash Medical Centre, Melbourne, Victoria; Women’s and Children’s Hospital, Adelaide, South Australia; and Perth Children’s Hospital, Perth, Western Australia: and Royal Darwin Hospital, Darwin, Northern Territory. Collectively, these states and territory include 96.1% of the Australian pediatric population. Children who were immunosuppressed or receiving intravenous immunoglobulin were excluded from the study as they may not mount a representative antibody response to infection or because immunoglobulin use could influence detection of SARS-CoV-2-specific antibodies. Participants were preferentially recruited from day stay lists of patients undergoing minor procedures to maximize recruitment of children without complex medical conditions. Written consent, on paper or the RedCap database was provided by parents/guardians of children aged <18 years and by those aged 18-19 years themselves. In addition, written assent was provided by adolescents aged ≥12 years. Blood collection occurred during intravenous cannulation following anesthetic induction. A questionnaire was administered to obtain demographic information (age, sex, Indigenous status, postcode of residence), history and timing of known SARS-CoV-2 infection, date of vaccination dose 1 and 2, and underlying medical conditions. An additional questionnaire was administered if infants were <1 year of age to ask about history of maternal SARS-CoV-2 infection and vaccination prior to delivery.

States and territories had differing public health measures and patterns of notified cases. We sought to obtain seroprevalence estimates for each jurisdiction separately, as well as a national estimate. A planned sample size of 385 samples in each jurisdiction was calculated based on a desired maximum 95% confidence interval (CI) width of +/-5%. Greater precision with a maximum 95% CI width of +/-3.6% would be achieved if the true population prevalence in the relevant subgroup was 85% or higher.

### Sample processing and testing

Blood samples were centrifuged at each site and sera were separated and extracted. The serum samples were coded and de-identified before sending to Victoria Infectious Diseases Reference Laboratory, Melbourne, Victoria for testing. Antibody testing was performed using the Roche Elecsys Anti-SARS-CoV-2 S-antibody and N-antibody electro-chemiluminescence immunoassays which detect antibodies of all immunoglobulin classes against the receptor-binding domain of the S-protein and the N-protein, respectively. Specimens were deemed positive for the Anti-SARS-CoV-2 N-assay if the semiquantitative cut-off index (COI) was ≥1.0, and for the Anti-SARS-CoV-2 S-assay if the quantitative result was ≥0.8 U/ml, as per the manufacturer’s instructions for use for each test. These assays have been used in pediatric serosurveys in England(18, 19) and Texas, USA(20). Anti-SARS-CoV-2 S-antibody results >250 IU/ml, underwent retesting after a 1:10 dilution to determine the quantitative result.

The manufacturer’s reported anti-S assay sensitivity was 98.8% (95%CI: 98.1%-99.3%) in individuals who were infected ≥14 days with the ancestral strain and specificity of 100% (95%CI: 99.7-100%). The anti-N assay sensitivity was 98.8% (95% CI 98.1-99.3) in individuals who were infected 2-35 weeks prior with the SARS-CoV-2 ancestral strain and specificity was 99.8% (95% CI 99.69 – 99.88)(21). Local validation studies using 252 samples from vaccinated 97 Australian health care workers found 89.7% (95% CI 85.3-92.9) seropositivity for N-antibody in samples collected between collected between 14 and 216 days post illness (unpublished). Assay specificity was assessed as 100% for both S-antibody and N-antibody when tested against a panel of pre-pandemic serum samples from Australian adults (unpublished). Similar high sensitivity and specificity have been elicited in several studies, in both adults and children(22–25).

### Data storage and analysis

Information on participants was entered into a Research Electronic Data Capture (REDCap^®^) online database, accessible by the respective PAEDS site in each state. Data linkage was performed to link serosurvey participants to PAEDS SARS-CoV-2 hospitalization data. Serological results were reported via email from the study team to the parents/carers of the participants.

De-identified information from the study database was extracted for analysis using Microsoft Excel, STATA, and R.

As well as calculating crude seroprevalence estimates of anti-SARS-CoV-2 antibodies, we applied multilevel regression and poststratification within a Bayesian framework (26) (27) to: (i) separately model the variation in SARS-CoV-2 S- and N-antibodies by the measured demographic covariates of age (0-6 months, 6-12 months, 1-4 years, 5-11 years, 12-15 years, 16-19 years), vaccination status (0, 1, ≥2-doses), socioeconomic quintile (as measured by ABS Socio-Economic Indexes for Areas 2016, Index of Relative Socio-economic Disadvantage), state or territory of residence and residential area of remoteness (major cities, regional, remote); and (ii) obtain prevalence estimates adjusted for imbalances with respect to these covariates in the participant sample relative to the national population aged 0–19 years. This was achieved by weighting model-based prevalence estimates for all possible combinations of these covariates by the corresponding covariate distribution in the population. Estimation assumed a uniform prior distribution for seroprevalence. We summarized seroprevalence estimates for the overall national population aged 0–19 years and various subgroups of interest using the median and 95% credible interval (CrI) of the corresponding posterior probability distribution. Adjusted estimates were not calculated for sex, Indigenous status, presence of co-morbidities, reported history of past infection as population data on these characteristics in combination with other covariates were not available.

Daily cumulative COVID-19 vaccination coverage (%) of doses 1 and 2 between January 1, 2021 and December 31, 2022 were estimated nationally by using the number of vaccinated people in each age group (5-11 years, 12-15 years and ≥16 years from the Australian Immunisation Register at April 2, 2023) as the numerator and the Australian Bureau of Statistics Estimated Resident Population (ABS-ERP) as of 30 June 2021 for each age group as the denominator. Infection fatality rate and infection hospital rate were calculated using data collected through the National Notifiable Diseases Surveillance System (NNDSS), population count from the ABS-ERP 30 June 2021 and adjusted S-antibody seroprevalence in unvaccinated children, determined through this serosurvey.

The S-antibody titers and N-antibody COI for both unvaccinated and 2-dose vaccinated participants were plotted by time from last vaccination or infection (whichever occurred more recently) to time of specimen collection. A locally weighted regression (loess) line was applied, and the overall median was calculated using R. The comparison between these medians was conducted using the Wilcoxon rank-sum test. Very few participants with reported infection had 1 or >2 vaccination doses and were not included.

### Ethics

Ethics approval for this national study was provided by the Sydney Children’s Hospital Network Human Research Ethics Committee (HREC 18/SCHN/72).

## Results

We obtained 2046 samples from 2314 consented participants (Fig 1). Of all samples 2045 were tested for S-antibody and N-antibody; 1 sample was tested solely for N-antibody due to insufficient volume. Blood collection occurred between June 1 and August 31, 2022, at a time of 73.5% and 38.4% 2-dose COVID-19 vaccine coverage in children aged 12-15 years and 5-11 years respectively (Fig 2). The demographic of those tested are described in Table 1: 872 (42.6%) participants were female and 177 (8.7%) were Aboriginal or Torres Strait Islander peoples. The majority (1315/2043, 64.4%) of children had no underlying medical conditions. Among those with an underlying medical condition, airway/chest disease (29.2%; 213/728), cardiac disease (13.9%; 101/728), and gastrointestinal disease (13.3%; 97/728) were most common (Supplementary Table 1). Past SARS-CoV-2 infection was reported in 49.9% (1014/2033) of participants and 57.8% (1179/2040) were unvaccinated, and varied by age. Participants came from all socioeconomic quintiles and most resided in metropolitan (73.1%; 1495/2046) regions, compared to regional (24.4%; 499/2046) and remote regions (2.5%, 52/2046) (Table 1). A small number of participants resided in the two other small jurisdictions not targeted in the study: Australian Capital Territory (n=21) and the state of Tasmania (n=14). Fig 3 shows the postcode of participants compared to the population geographic distribution of Australia.

**Fig 1:**
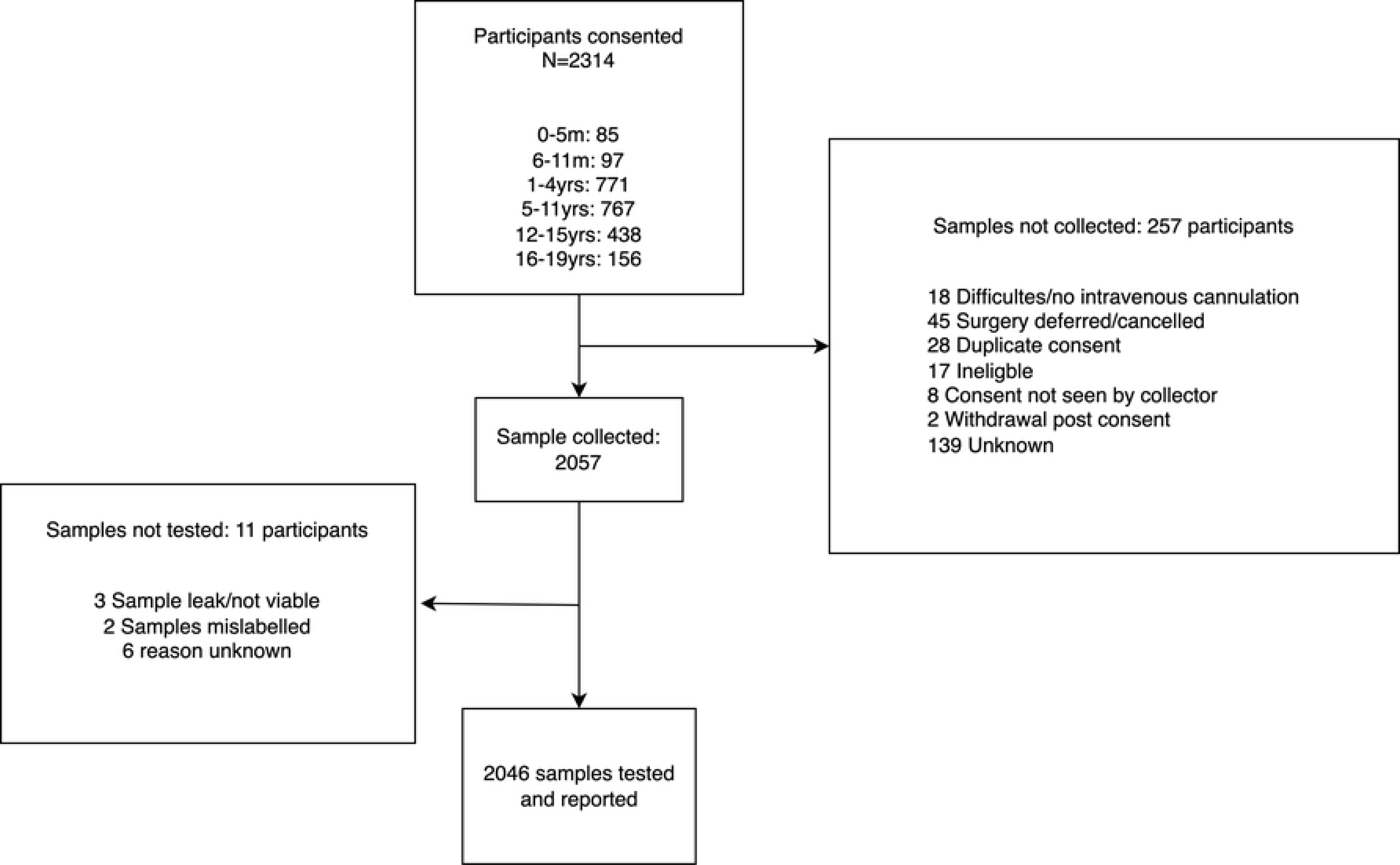
Participant recruitment and sample collection of children and adolescents aged 0-19 years for SARS-CoV-2 spike and nucleocapsid antibodies while undergoing an anaesthetic procedure in a PAEDS hospital, between June 1 and August 31, 2022 in Australia.

**Fig 2:**
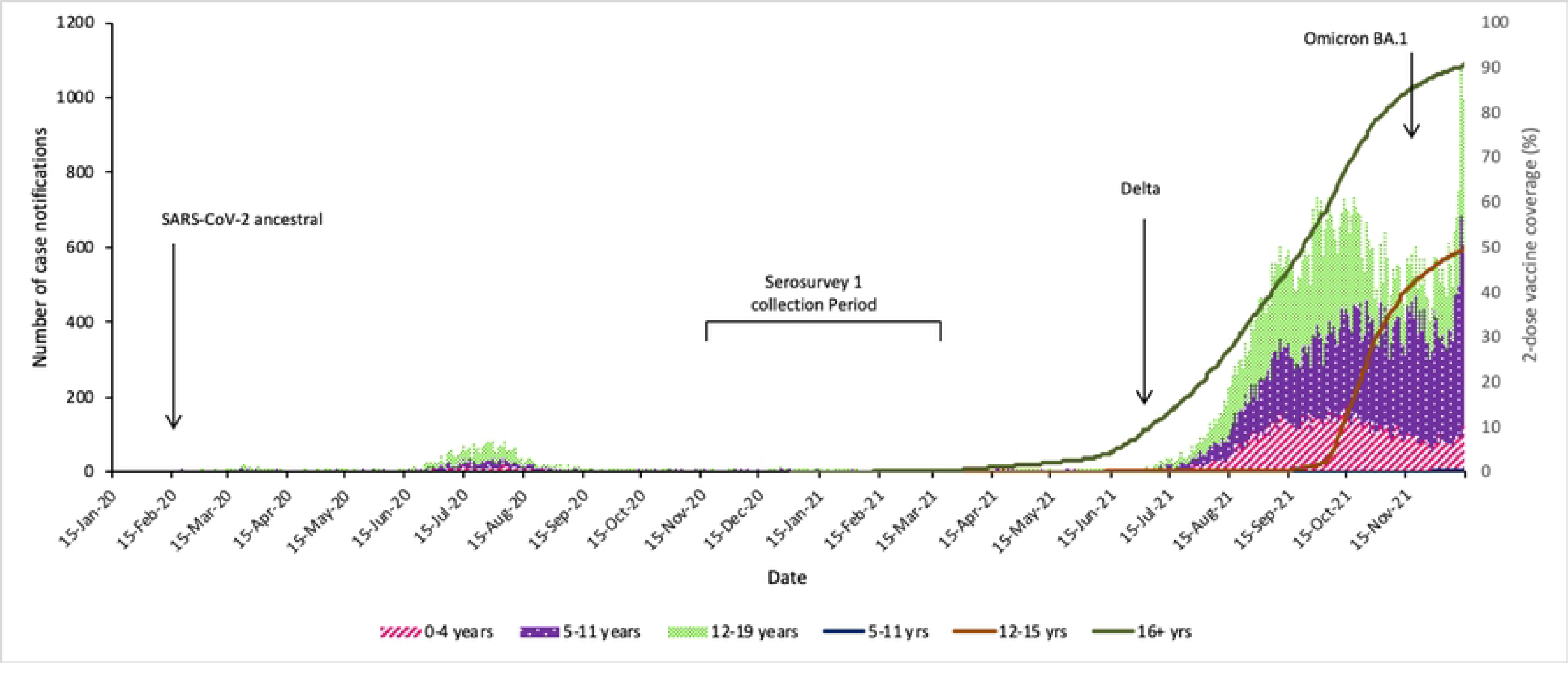
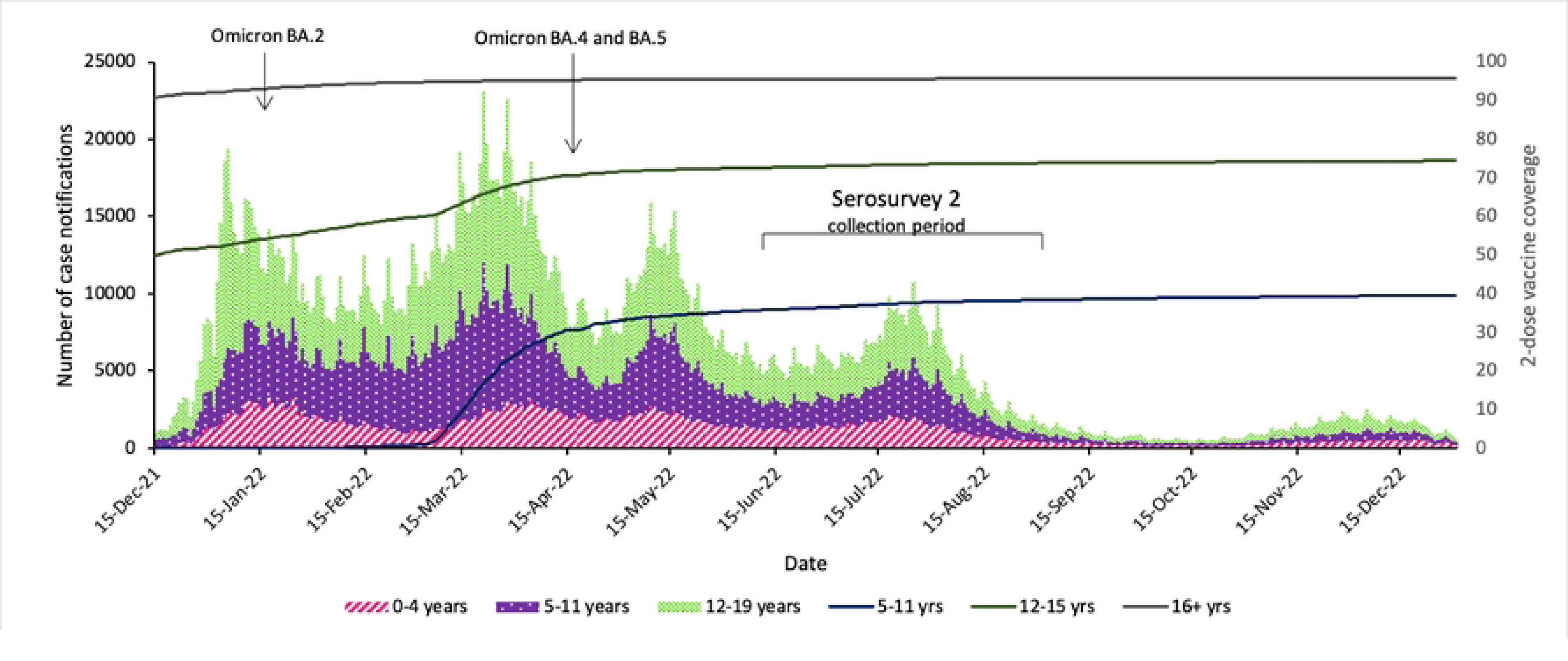
Number of SARS-CoV-2 infection notifications (bars) by calendar week in children aged 0-19 years in two distinct periods: a) from January 2020-November 2021 (the pre-Omicron era and period of strict public health measures, including international and internal border closures) and b) December 2021 – December 2022 (post Omicron period and with progressive easing of public health measures), and high levels of population vaccination coverage in adults and adolescents (lines; %).

**Figure 3:**
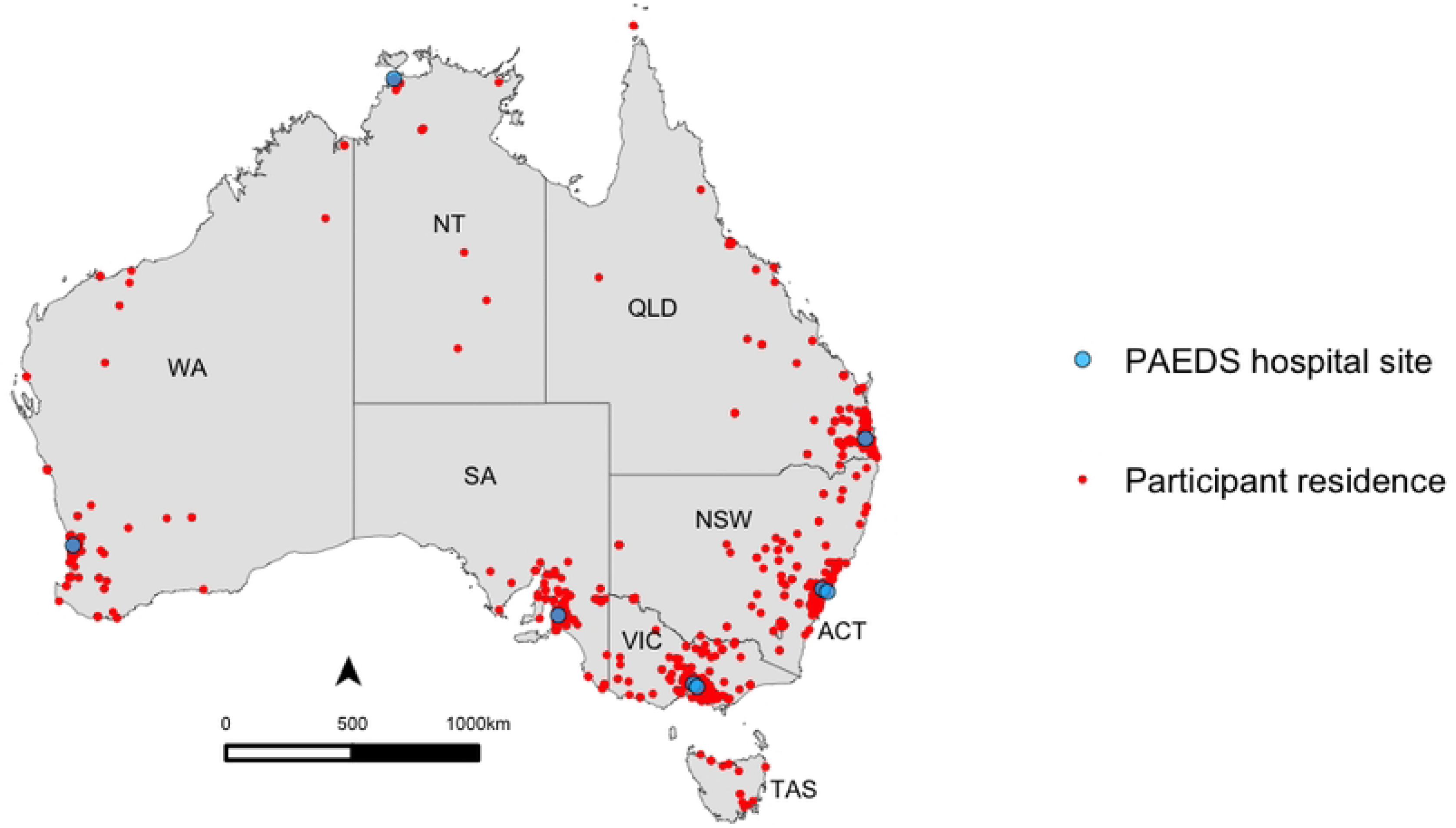
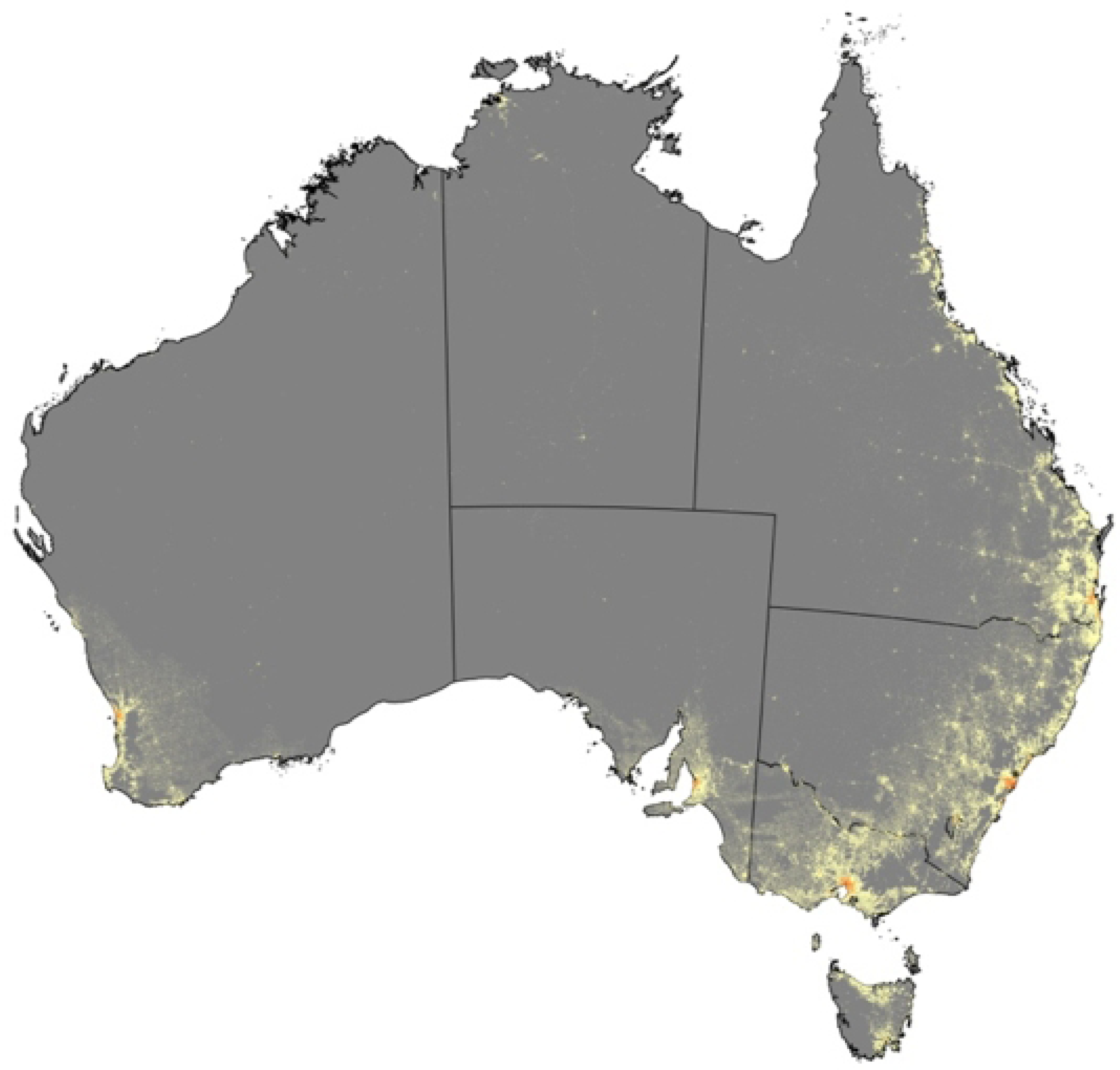
Map of a) postcode of residence^a^ of participants aged 0-19 years tested for SARS-CoV-2 spike and nucleocapsid antibodies while undergoing an anaesthetic procedure in a PAEDS hospital, between June 1 and August 31, 2023, in Australia compared to b) population geographic distribution of Australia in 2022; Australian Bureau of Statistics ***Footnote*** Fig 3: ^a^ The dots represent the centromere of the postcodes and do not indicate the exact location of the participant’s residence. In the NT, to limit identification of participants in remote communities, participants residing within remote communities (n=16) have been removed from the map. NSW: New South Wales, QLD: Queensland, NT: Northern Territory, WA: Western Australia, SA: South Australia; VIC: Victoria, PCH: Perth Children’s Hospital, WCH: Women’s Children’s Hospital.

**Table 1:**
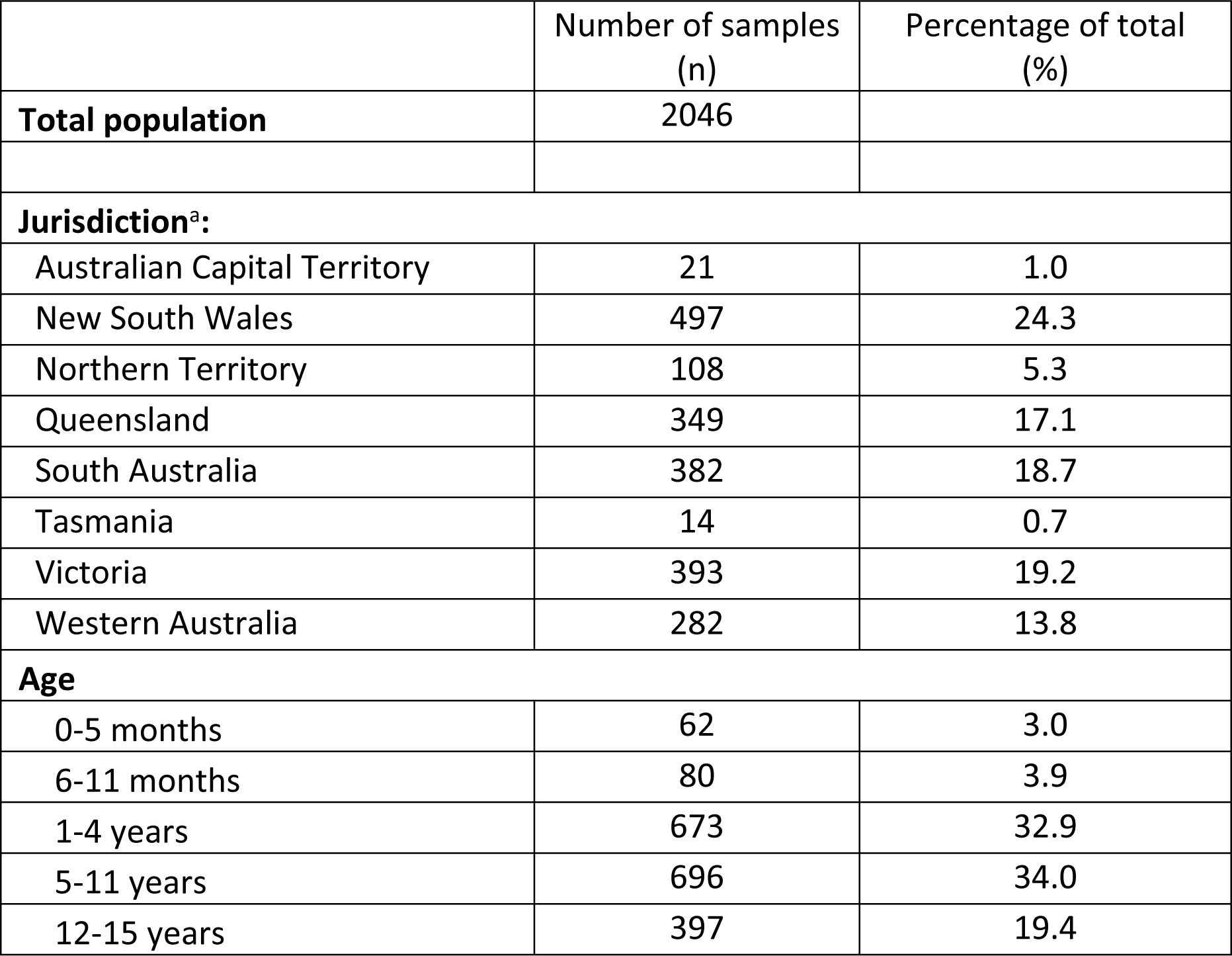
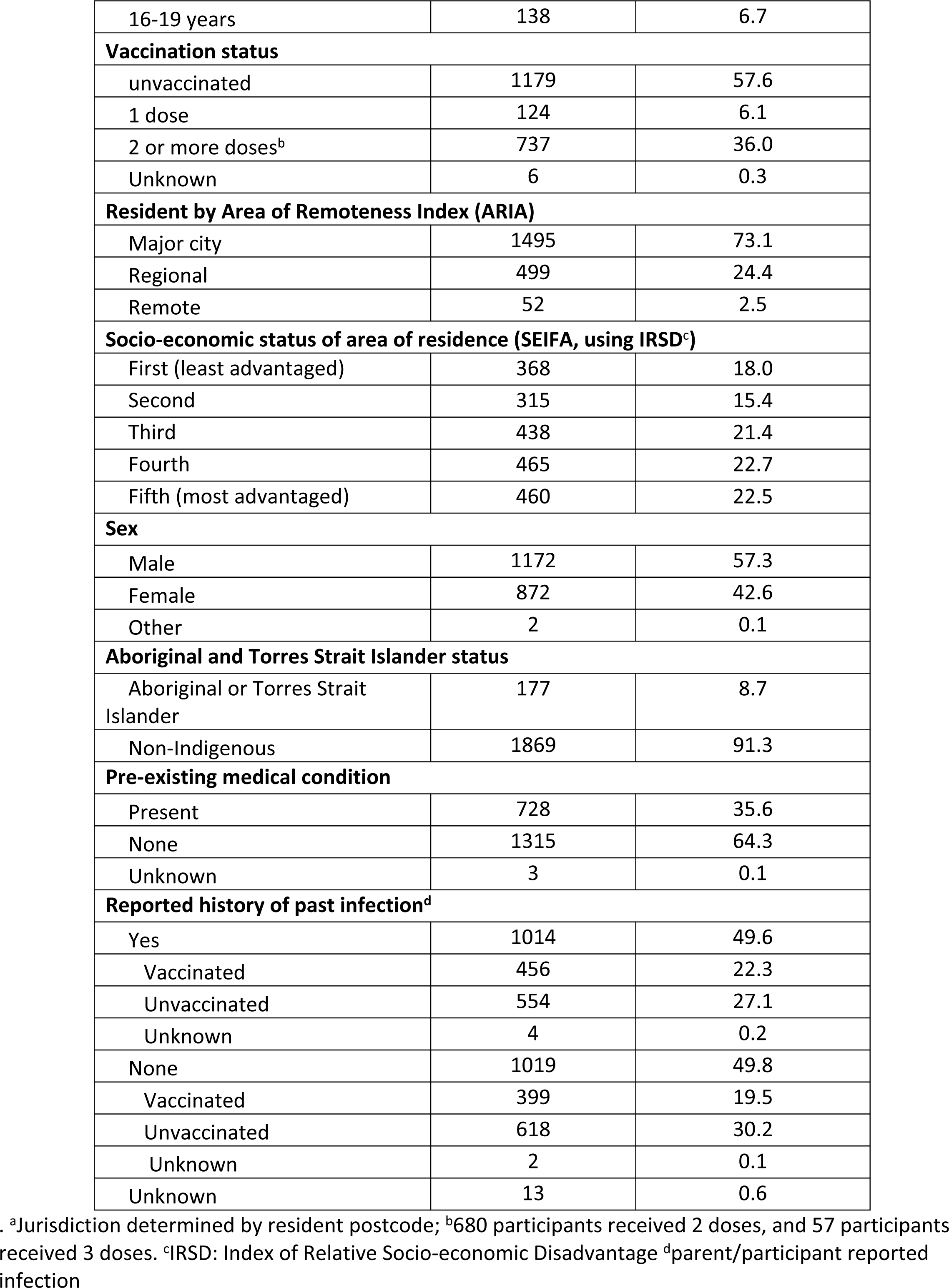
Demographics of children and adolescents aged 0-19 years tested for SARS-CoV-2 spike and nucleocapsid antibodies while undergoing an anesthetic procedure in a PAEDS hospital, between June 1 and August 31, 2022, in Australia.

|  | Number of samples<br>(n) | Percentage of total<br>(%) |
| --- | --- | --- |
| **Total population** | 2046 |  |
| **Jurisdiction<sup>a</sup>:** |  |  |
| Australian Capital Territory | 21 | 1.0 |
| New South Wales | 497 | 24.3 |
| Northern Territory | 108 | 5.3 |
| Queensland | 349 | 17.1 |
| South Australia | 382 | 18.7 |
| Tasmania | 14 | 0.7 |
| Victoria | 393 | 19.2 |
| Western Australia | 282 | 13.8 |
| **Age** |  |  |
| 0-5 months | 62 | 3.0 |
| 6-11 months | 80 | 3.9 |
| 1-4 years | 673 | 32.9 |
| 5-11 years | 696 | 34.0 |
| 12-15 years | 397 | 19.4 |
| 16-19 years | 138 | 6.7 |
| **Vaccination status** |  |  |
| unvaccinated | 1179 | 57.6 |
| 1 dose | 124 | 6.1 |
| 2 or more doses<sup>b</sup> | 737 | 36.0 |
| Unknown | 6 | 0.3 |
| **Resident by Area of Remoteness Index (ARIA)** |  |  |
| Major city | 1495 | 73.1 |
| Regional | 499 | 24.4 |
| Remote | 52 | 2.5 |
| **Socio-economic status of area of residence (SEIFA, using IRSD<sup>c</sup>)** |  |  |
| First (least advantaged) | 368 | 18.0 |
| Second | 315 | 15.4 |
| Third | 438 | 21.4 |
| Fourth | 465 | 22.7 |
| Fifth (most advantaged) | 460 | 22.5 |
| **Sex** |  |  |
| Male | 1172 | 57.3 |
| Female | 872 | 42.6 |
| Other | 2 | 0.1 |
| **Aboriginal and Torres Strait Islander status** |  |  |
| Aboriginal or Torres Strait Islander | 177 | 8.7 |
| Non-Indigenous | 1869 | 91.3 |
| **Pre-existing medical condition** |  |  |
| Present | 728 | 35.6 |
| None | 1315 | 64.3 |
| Unknown | 3 | 0.1 |
| **Reported history of past infection<sup>d</sup>** |  |  |
| Yes | 1014 | 49.6 |
| Vaccinated | 456 | 22.3 |
| Unvaccinated | 554 | 27.1 |
| Unknown | 4 | 0.2 |
| None | 1019 | 49.8 |
| Vaccinated | 399 | 19.5 |
| Unvaccinated | 618 | 30.2 |
| Unknown | 2 | 0.1 |
| Unknown | 13 | 0.6 |

. <sup>a</sup>Jurisdiction determined by resident postcode; <sup>b</sup>680 participants received 2 doses, and 57 participants received 3 doses. <sup>c</sup>IRSD: Index of Relative Socio-economic Disadvantage <sup>d</sup>parent/participant reported infection

Crude seroprevalence of S-antibody was 1833/2045 (89.6%) and N-antibody 1309/2046 (64.0%). After adjustment for age, vaccination status, socioeconomic quintile and remoteness index, seroprevalence for S-antibody was 92.1% (95% CrI 91.0%-93.3%) and N-antibody 67.0% (95% CrI 64.6%-69.3%).

Seroprevalence of both S-antibody and N-antibody increased with age but was similar across jurisdictions and socioeconomic quintiles (Fig 4). S-antibody was present in all children who received 1 or ≥2 doses of the COVID-19 vaccine and an adjusted estimate of 84.2% (95% CrI 81.9-86.5) of unvaccinated participants. N-antibody was detected at similar levels among ≥2-dose vaccinated (adjusted estimate of 66.5%; 95%CrI 62.9-70.4), 1-dose vaccinated (adjusted estimate of 70.7%; 95% CrI 63.7-77.7) and unvaccinated individuals (adjusted estimate of 67.1%; 95%CrI 64.0%-69.8%).

**Fig 4:**
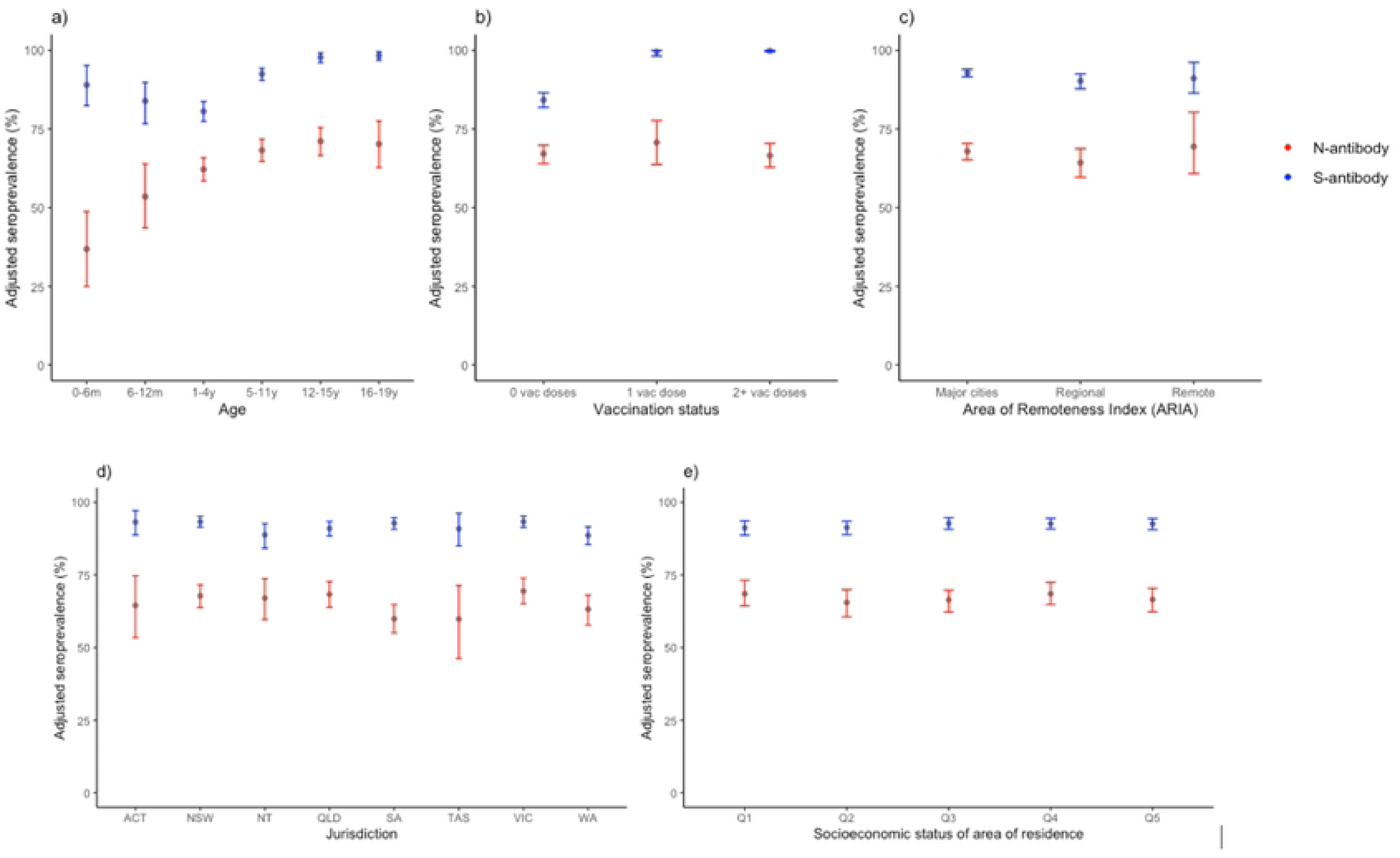
Adjusted seroprevalence estimates for SARS-CoV-2 spike antibody (S-antibody) and nucleocapsid antibody (N-antibody) collected from children and adolescents aged 0-19 years undergoing anaesthetic procedures between June 1 and August 31, 2022 by: a) age, b) vaccination status, c) area of remoteness Index (ARIA), d) jurisdiction, and e) socio-economic status of area of residence ***Footnote*** Fig 4: ACT: Australian Capital Territory, NSW: New South Wales, NT: Northern Territory, QLD: Queensland, SA: South Australia, TAS: Tasmania, VIC: Victoria, WA: Western Australia; SEIFA quintile: measured using the Index of Relative Socio-economic Disadvantage; Q1 – highest quintile, and Q5 – lowest quintile;

In participants who were unvaccinated and reported a past infection, the crude S-antibody positivity was 96.2% (522/554) and crude N-antibody positivity was 88.2% (489/554). In participants who were vaccinated and reported infection, the crude S-antibody positivity was 100% (455/455) and the crude N-antibody positivity was 420/455 (92.3%) (Table 2). S and N-antibody positivity among children with 1 or more underlying medical conditions were 88.6% (644/727) and 59.1% (730/728) respectively. A breakdown of seroprevalence by medical conditions can be found in Supplementary Table 1.

**Table 2:** Seroprevalence of SARS-CoV-2 spike antibody (S-antibody) and nucleocapsid antibody (N-antibody) estimated from blood samples taken from children and young adults aged 0-19 years undergoing an anesthetic procedure in a PAEDS hospital, in Australia between June 1 and August 31, 2022, by participant demographics, socio-economic status of participant residential postcode (Socio-Economic Index for Areas, SEIFA; Index of Relative Socio-economic Disadvantage, IRSD) by quintile, geographic classification of participant residential postcode (Accessibility Remoteness Index of Australia, ARIA).

|  | S- antibody n/N(%) |  | N-antibody n/N(%) |  |
| --- | --- | --- | --- | --- |
|  | Crude n/N (%) | Adjusted<sup>a</sup> % (95% credible interval) | Crude n/N (%) | Adjusted<sup>a</sup> % (95% credible interval) |
| **Total population** | 1833/2045 (89.6) | 92.1 (91.0 - 93.3) | 1309/2046 (64.0) | 67.0 (64.6 - 69.3) |
| **Jurisdiction<sup>b</sup>:** |  |  |  |  |
| Australian Capital Territory | 19/21 (90.5) | 93.2 (88.8 - 97.1) | 13/21 (61.9) | 64.5 (53.4 - 74.6) |
| New South Wales | 458/497 (92.2) | 93.3 (91.5 - 95.1) | 329/497 (66.2) | 67.8 (63.8 - 71.6) |
| Northern Territory | 90/108 (83.3) | 88.8 (84.2 - 92.7) | 71/108 (65.7) | 67.0 (59.6 - 73.7) |
| Queensland | 314/349 (90.0) | 91.0 (88.4 - 93.4) | 236/349 (67.6) | 68.3 (63.9 - 72.7) |
| South Australia | 348/381 (91.3) | 92.8 (90.7 - 94.7) | 216/382 (56.5) | 59.9 (55.0 - 64.8) |
| Tasmania | 13/14 (92.9) | 90.9 (85.0 - 96.2) | 6/14 (42.9) | 59.8 (46.2 - 71.3) |
| Victoria | 361/393 (91.9) | 93.3 (91.4 - 95.2) | 271/393 (69.0) | 69.4 (65.1 - 73.9) |
| Western Australia | 230/282 (81.6) | 88.6 (85.5 - 91.6) | 167/282 (59.2) | 63.2 (57.7 - 68.0) |
| **Age** |  |  |  |  |
| 0-5 months | 57/62 (91.9) | 89.0 (82.4 - 95.2) | 20/62 (32.3) | 36.8 (25.0 - 48.7) |
| 6-11 months | 65/80 (81.3) | 83.9 (76.7 - 89.8) | 41/80 (51.3) | 53.5 (43.5 - 63.9) |
| 1-4 years | 531/673 (78.9) | 80.6 (77.4 - 83.7) | 409/673 (60.8) | 62.2 (58.5 - 65.8) |
| 5-11 years | 650/696 (93.4) | 92.5 (90.5 - 94.3) | 465/696 (66.8) | 68.2 (64.7 - 71.7) |
| 12-15 years | 394/397 (99.2) | 97.8 (96.1 - 99.2) | 278/397 (70.0) | 71.1 (66.5 - 75.4) |
| 16-19 years | 136/137 (99.3) | 98.3 (96.9 - 99.5) | 96/138 (69.6) | 70.2 (62.8 - 77.5) |
| **Vaccination status** |  |  |  |  |
| unvaccinated | 967/1179 (82.0) | 84.2 (81.9 - 86.5) | 743/1179 (63.0) | 67.1 (64.0 - 69.8) |
| 1 dose | 124/124 (100.0) | 99.5 (98.2 - 100) | 86/124 (69.4) | 70.7 (63.7 - 77.7) |
| 2 or more doses<sup>c</sup> | 736/736 (100.0) | 99.8 (99.5 - 100) | 475/737 (64.5) | 66.5 (62.9 - 70.4) |
| **Resident by Area of Remoteness Index (ARIA)** |  |  |  |  |
| Major city | 1356/1495 (90.7) | 92.8 (91.6 - 94.0) | 969/1495 (64.8) | 67.9 (65.2 - 70.4) |
| Regional | 429/498 (86.1) | 90.3 (87.8 - 92.5) | 302/499 (60.5) | 64.3 (59.7 - 68.7) |
| Remote | 48/52 (92.3) | 91.1 (86.4 - 96.2) | 38/52 (73.1) | 69.4 (60.9 - 80.3) |
| **Socio-economic status of area of residence (SEIFA, using IRSD<sup>d</sup>)** |  |  |  |  |
| First (least advantaged) | 328/368 (89.1) | 91.2 (88.7 - 93.6) | 244/368 (66.3) | 68.5 (64.3 - 73.2) |
| Second | 280/315 (88.9) | 91.3 (88.9 - 93.5) | 190/315 (60.3) | 65.5 (60.6 - 69.9) |
| Third | 399/437 (91.3) | 92.7 (90.7 - 94.7) | 274/438 (62.6) | 66.4 (62.2 - 69.7) |
| Fourth | 422/465 (90.8) | 92.6 (90.8 - 94.5) | 311/465 (66.9) | 68.5 (64.8 - 72.4) |
| Fifth (most advantaged) | 404/460 (87.8) | 92.6 (90.6 - 94.4) | 290/460 (63.0) | 66.5 (62.3 - 70.4) |
| **Sex** |  |  |  |  |
| Male | 1039/1171 (88.7) | - | 732/1172 (62.5) | - |
| Female | 792/872 (90.8) | - | 576/872 (66.1) | - |
| Other | 2/2 (100.0) | - | 1/2 (50.0) | - |
| **Aboriginal and Torres Strait Islander status** |  |  |  |  |
| Aboriginal and Torres Strait Islander | 159/177 (89.8) | - | 115/177 (65.0) | - |
| Non-Indigenous | 1674/1868 (89.6) | - | 1194/1869 (63.9) | - |
| **Underlying medical conditions** |  |  |  |  |
| Present | 644/727 (88.6) | - | 430/728 (59.1) | - |
| None | 1187/1315 (90.3) | - | 878/1315 (66.8) | - |
| **Reported history of past infection** |  |  |  |  |
| Yes | 992/1013 (97.9) | - | 914/1014 (90.1) | - |
| Vaccinated | 455/455 (100%) |  | 420/455 (92.3%) |  |
| Unvaccinated | 533/554 (96.2%) |  | 489/554 (88.3%) |  |
| None | 831/1019 (81.6) | - | 387/1019 (38.0) | - |
| Vaccinated | 399/399 (100%) |  | 136/399 (34.1%) |  |
| Unvaccinated | 430/618 (69.6%) |  | 250/618 (40.4%) |  |

<sup>a</sup>Adjusted estimates are not provided for sex, Aboriginal and Torres Strait Islander status, presence of co-morbidities, reported history of past infection as population data on these characteristics in combination with other covariates were not available. <sup>b</sup>Jurisdiction determined by resident postcode; <sup>c</sup>680 participants received 2 doses, and 57 participants received 3 doses. <sup>d</sup>IRSD: Index of Relative Socio-economic Disadvantage

In infants aged 0-11 months, S-antibody seroprevalence was high: adjusted estimates in those aged 0-5 months was 89.0% (95% CrI 82.4%-95.2%) and among those aged 6-11 months it was 83.9% (95% CrI 76.7-89.3). The adjusted N-antibody seroprevalence estimate increased from 36.8% (95% CrI 25.0%-48.7%) at 0-5 months to 53.5% (95% CrI 43.5%-63.9%) at 6-11 months (Table 2). Data on past maternal infection and vaccination were available in 107/142 (75.4%) infants aged 0-11 months. S-antibody was detected universally (48/48) in infants aged 0-5 months in mothers who had been vaccinated, regardless of history of prior infant infection. In infants aged 6-11 months, S-antibody was detected in 15/15 infants who had reported prior infection but only in 15/17 infants with no reported prior infection. Seroprevalence of S-and N-antibody within this population is shown on Table 3.

**Table 3:**
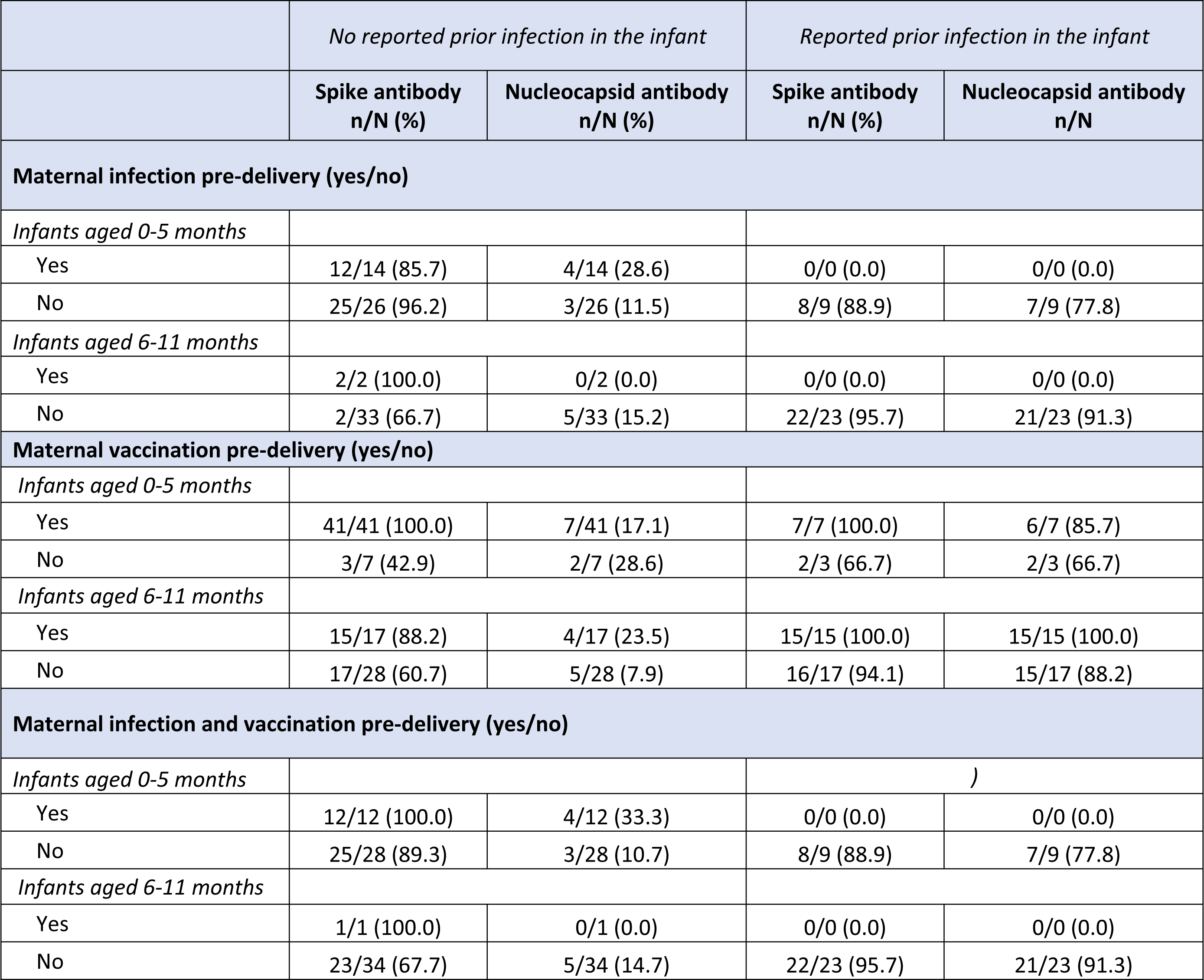
Seroprevalence of SARS-CoV-2 spike and nucleocapsid antibody estimated from blood samples taken from infants aged 0-5 months and 6-11 months by maternal infection and/or vaccination status prior to delivery.

|  | *No reported prior infection in the infant* |  | *Reported prior infection in the infant* |  |
| --- | --- | --- | --- | --- |
|  | **Spike antibody**<br>**n/N (%)** | **Nucleocapsid antibody**<br>**n/N (%)** | **Spike antibody**<br>**n/N (%)** | **Nucleocapsid antibody**<br>**n/N** |
| **Maternal infection pre-delivery (yes/no)** |  |  |  |  |
| *Infants aged 0-5 months* |  |  |  |  |
| Yes | 12/14 (85.7) | 4/14 (28.6) | 0/0 (0.0) | 0/0 (0.0) |
| No | 25/26 (96.2) | 3/26 (11.5) | 8/9 (88.9) | 7/9 (77.8) |
| *Infants aged 6-11 months* |  |  |  |  |
| Yes | 2/2 (100.0) | 0/2 (0.0) | 0/0 (0.0) | 0/0 (0.0) |
| No | 2/33 (66.7) | 5/33 (15.2) | 22/23 (95.7) | 21/23 (91.3) |
| **Maternal vaccination pre-delivery (yes/no)** |  |  |  |  |
| *Infants aged 0-5 months* |  |  |  |  |
| Yes | 41/41 (100.0) | 7/41 (17.1) | 7/7 (100.0) | 6/7 (85.7) |
| No | 3/7 (42.9) | 2/7 (28.6) | 2/3 (66.7) | 2/3 (66.7) |
| *Infants aged 6-11 months* |  |  |  |  |
| Yes | 15/17 (88.2) | 4/17 (23.5) | 15/15 (100.0) | 15/15 (100.0) |
| No | 17/28 (60.7) | 5/28 (7.9) | 16/17 (94.1) | 15/17 (88.2) |
| **Maternal infection and vaccination pre-delivery (yes/no)** |  |  |  |  |
| *Infants aged 0-5 months* |  |  |  | ) |
| Yes | 12/12 (100.0) | 4/12 (33.3) | 0/0 (0.0) | 0/0 (0.0) |
| No | 25/28 (89.3) | 3/28 (10.7) | 8/9 (88.9) | 7/9 (77.8) |
| *Infants aged 6-11 months* |  |  |  |  |
| Yes | 1/1 (100.0) | 0/1 (0.0) | 0/0 (0.0) | 0/0 (0.0) |
| No | 23/34 (67.7) | 5/34 (14.7) | 22/23 (95.7) | 21/23 (91.3) |

Participants who reported a past infection and had received 2-doses of a COVID-19 vaccine had a median S-antibody titer of 12318 U/mL which was significantly (349 U/mL-fold, p<0.001) higher than the median S-antibody titer of 35.3 u/L of unvaccinated participants who reported a past infection. No significant difference was detected in the median N-antibody COI (19.0 in 2-dose vaccinated, 16.3 in the unvaccinated, difference 1.16; p=0.55) between the two groups. Fig 5 shows there was minimal waning of S-antibody within 300 days in both groups. N-antibody showed the natural rise in COI for the first 14 days and also remained stable within 300 days.

**Fig 5.**
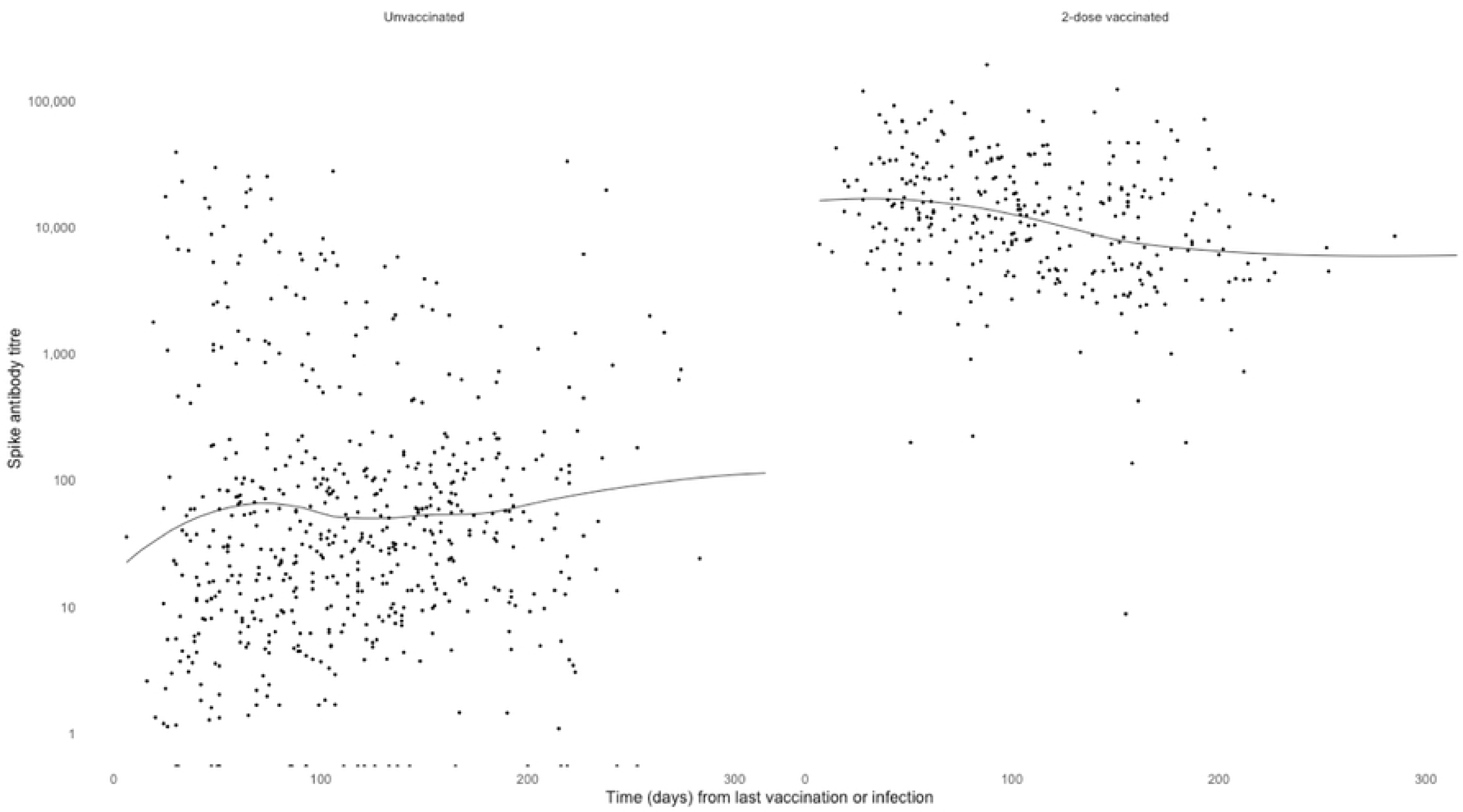
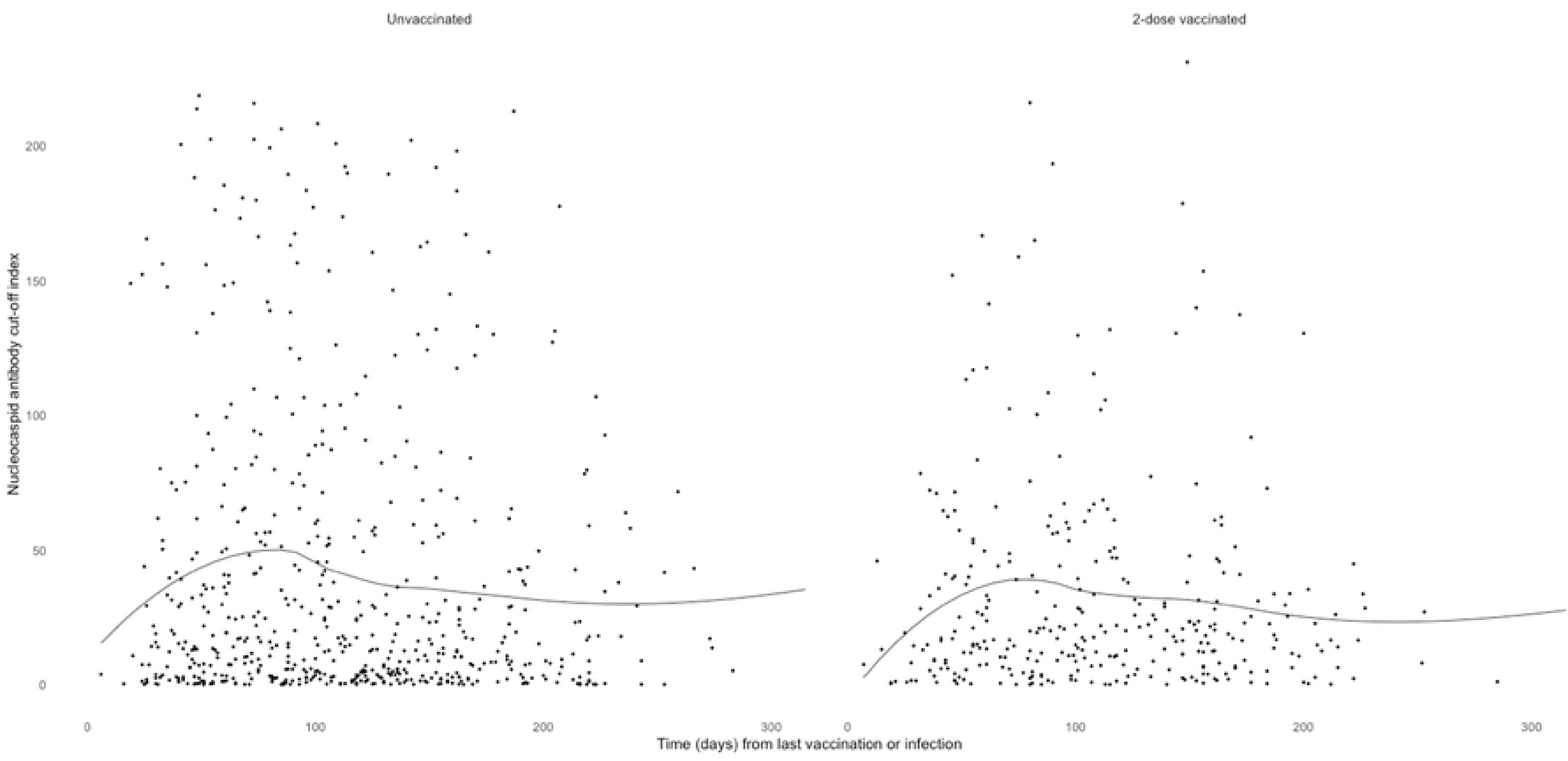
a) SARS-CoV-2 spike antibody titer and b) nucleocapsid antibody cut-off index from blood samples from participants aged 0-19 years with a prior history of SARS-CoV-2 infection by time (days) from last vaccination or infection and vaccination status^a^. ***Footnote*** Fig 5: ^a^Locally weight regression (loess) was utilized to create a line of best fit on R.

In the Australian National Notifiable Diseases Surveillance System dataset, between 25 January 2020 and 31 August 2022, there were 2,236,528 COVID-19 case notifications among children and adolescents aged 0-19 years. Of these 16,358 (0.7%) were hospitalized and or admitted to an Emergency Department with SARS-CoV-2 infection and 26 were reported to have died with COVID-19;(28) using Australian Bureau of Statistics (ABS) quarterly population estimate for June 2022(29) this equated to a crude case fatality rate of 0.01 per 1000). Alternatively, using cumulative adjusted S-antibody seroprevalence rates for unvaccinated children calculated through this serosurvey as the infection rate, the SARS-CoV-2 infection hospitalization rate (including emergency department encounters) was 307 per 100,000 (95% CrI 299 – 316 per 100,000 infections) and the infection fatality rate was 0.0049 per 1,000 infections (95% CrI 0.0048-0.0050 per 1,000) in those aged 0-19 years.

Only 23/2046 serosurvey participants (1.1%) had been admitted at one of the PAEDS sentinel hospitals with SARS-CoV-2 infection prior to participating in the serosurvey. Most had underlying medical conditions (19/23; 82.6%). The median age was 1.9 years (age range 2 days to 15.6 years). Twelve were admitted for fever and respiratory distress (COVID-19), 2 were admitted for unrelated bone fractures, 3 for seizures, 1 for management of chronic constipation and 1 for new diagnosis of Type 1 diabetes mellitus. Four children required intensive care admission – 3 for acute SARS-CoV-2 pneumonitis, of which 2 required invasive ventilation and 1 for diabetic ketoacidosis. All 3 children admitted to ICU with COVID pneumonitis had either complex genetic conditions and/ or underlying cardiorespiratory disease.

## Discussion

In Australia by August 2022, the estimated seroprevalence in children and adolescents of SARS-CoV-2 Spike antibody was 92.1% (95% CI 91% - 93.3%), and N-antibody was 67% (95% CI 64.6% - 69.3%). Thereby indicating the majority of those aged 0-19 years had been infected with SARS-CoV-2. Based on prevalence of S-antibody in unvaccinated children, data (indicative of infection) the true infection rate may be as high as 80%, estimating that over 5 million Australian children have been infected by August 31, 2022. This represents a large increase from August 2021 when <1% of the pediatric population had been infected.(13) Infection rates were across states and territories, metropolitan, regional and rural geographic regions and across socioeconomic quintiles, suggesting that Omicron variant SARS-CoV-2 infection spread rapidly and uniformly across Australia in a very short period of time. There was a higher rate of S-antibody and N-antibody positivity with increasing age and adults were similar to rates seen in adult blood donors (17). This is likely reflective of higher vaccination coverage and increased infection related to social mixing patterns in adolescents compared to primary school age children.

Despite high infection rates in children, using national notification data, we estimated the infection fatality and hospitalization rates to be very low at 0.0049 per 1,000 (95% CrI 0.0048-0.0050 per 1,000) and 307 per 100,000 (95% CrI 299 – 316 per 100,000). The rate of hospitalizations caused by COVID-19 may also be much lower as studies including our study have found many children are hospitalized for alternate reasons, whilst having concurrent SARS-CoV-2 infection. (30) (10) Our study also shows that majority of children under 5 years, who were ineligible for vaccination at the time of sera collection already had evidence of prior infection. However, S-antibody levels were higher in children who had been infected and vaccinated (hybrid immunity) compared to those only infected. In adults, hybrid immunity has been shown to protect against subsequent symptomatic infection and severe disease(31). The clinical benefit of immunization in children with low population infection fatality and infection hospitalization rates warrants longitudinal follow up surveillance and studies to better understand who is most at risk (e.g. due to underlying medical conditions), optimal vaccine schedules and better characterization of immunity, including non-humoral immunity overtime with repeated infection, in this population(32).

In infants < 6 months of age, our finding of higher S-antibody positivity (89%; 95% CrI 82.4-95.2) compared to 6-11 months in infants (83.9; 95%CrI 76.7-89.8) not reported to have had infection, suggests detected S-antibody was likely to have been maternal in origin. Longitudinal studies following mother and infant pairs have shown high rates of detection of SARS-CoV-2 antibodies in infants of vaccinated mothers, with rapid decline in maternally-derived antibodies from 6 months of age(33, 34) not dissimilar to antibody waning in infants whose mothers have been vaccinated in pregnancy for influenza.(35, 36) Although overall rates in infants are lower compared to adults, they face the highest rates of hospitalization within the pediatric age group.(37) (38) (39) Transplacental transfer of antibody to the fetus in pregnancy may prevent severe disease in young infants, emphasizing the importance of vaccinating pregnant women.(34, 39) Protection of infants < 6 months of age will be of ongoing interest with any new SARS CoV2 variants of concern, with seroprevalence studies helping to support the recommendations for vaccination of pregnant women.

We found using N-antibody alone may underestimate infection rates. In unvaccinated 1–19-year-olds in our cohort who reported past infection based on virus detection, crude S-antibody positivity was 96.2% (522/554) compared to crude N-antibody positivity of 88.2% (489/554). Compared to N-antibodies, S-antibodies are more persistent(40) but lack the ability to differentiate between infection and vaccination. In those who are vaccinated, S antibodies display a reduced magnitude of response and a quicker decline compared to their unvaccinated counterparts(41). Notably, in children with reported past infections, although S-antibody levels were significantly higher in those who had received 2 vaccine doses compared with those unvaccinated, the N-antibody COI response was similar in both groups, up to 8 months after infection. However, this observation comes from a cross-sectional study, limiting the ability to track individual variations over time post-vaccination or infection, thus preventing conclusive inferences about antibody levels and durability. A longitudinal school study of 184 infected participants (46 students and 138 staff members) in England, demonstrated that N-antibodies seroreversion occurred in 58.4% compared to S-antibody seroreversion of 20.9% by 24 weeks(42) and a study of 38 health care workers the geometry means of N-antibody COI declined from 77 (56.4-105) at 2 months post infection to 22.2 (13.1-37.9) at 18 months(43).

The strengths of this study were obtaining sera from children across Australia’s vast geographic range and all socioeconomic quintiles. The samples were broadly representative of the Australian pediatric population.(44) We were able to obtain some insights into S and N-antibody kinetics in vaccinated and unvaccinated children due to our large cohort of children and detailed data on infection and vaccination. Nevertheless, our study was necessarily pragmatic and cross-sectional in design and therefore had several limitations. These included opportunistic sampling which resulted a greater preponderance of males, however, sex is not a known risk factor for infection in children and thus unlikely to have biased seroprevalence levels. Reporting of past infection was based on recall of a positive laboratory test for SARS-CoV-2 and may be subject to biases related to recall and selective/under-testing in context of mild or no symptoms and more. The proportion of participants with an underlying medical condition in our survey was 35.6% (728/2043) which was lower than the ABS National Health Survey 2022 estimate of proportions of Australians aged 0-17 years with 1 or more current long-term health conditions (49.8% 95% CI 47.8%-52%).(45) Nevertheless, we were able to show that most children with underlying medical conditions have been previously infected, without severe disease. We had low numbers of older adolescents aged 16-19 years which was related to patterns of care as most in this age group receive care/procedures in adult hospitals. We also did not test the samples for neutralizing antibodies against circulating variants and so were unable to further characterize the antibody to determine the SARS-CoV-2 Omicron subvariant responsible for infection; future studies are being undertaken in this regard.

## Conclusion

In summary, by August 2022 most Australian children, spanning all geographic regions and socioeconomic quintiles had been infected with SARS-CoV-2. This indicates a swift and consistent spread of the virus throughout Australia within a brief timeframe to inform adaptive public health measures and vaccination strategies nationwide. Future data on seroprevalence should be complemented with studies of immune responses and correlated with disease outcomes to better understand infection in children and adolescents.

## Data Availability

All relevant data are within the manuscript and its Supporting Information files. Line listed data cannot be shared publicly because of identification of Aboriginal and Torres Strait Islander and non-Indigenous children and adolescents in regional and remote Australian settings, in particular participants from remote communities.

## Acknowledgements

Heather Gidding, Bette Liu, Mehyar Khair Baik, Deepika Jindal, Hayley Giuliano, Chelsea Bartel, Audrey Rattray, Jaimee Craft, Jill Nguyen, Klara Glavacevic, Mark Mayo, Vanessa Rigas, Celeste Woerle, Gemma Chamberlain, Margaret Lyon, Kelly McCrory, Karen Bellamy, Megan Wieringa, Joseph Speekman, Janine Maloney, Hilde Wegter, Annalisa Shine, Madison Bellamy, Melitta Allen, Michelle (Jia Xi) Li, Mai Khuu, Emma Leighton, Carolyn Pardo Vaccine Immunology Group, MCRI (Nadia Mazarakis, Rachel Higgins, Zheng Quan Toh

## References

1. Top KA, Macartney K, Bettinger JA, Tan B, Blyth CC, Marshall HS, et al. Active surveillance of acute paediatric hospitalisations demonstrates the impact of vaccination programmes and informs vaccine policy in Canada and Australia. Eurosurveillance. 2020;25(25):1900562.

2. Arora RK, Joseph A, Van Wyk J, Rocco S, Atmaja A, May E, et al. SeroTracker: a global SARS-CoV-2 seroprevalence dashboard. Lancet Infect Dis. 2021;21(4):e75–e6.

3. Bailey LC, Razzaghi H, Burrows EK, Bunnell HT, Camacho PEF, Christakis DA, et al. Assessment of 135 794 Pediatric Patients Tested for Severe Acute Respiratory Syndrome Coronavirus 2 Across the United States. Jama, Pediatr. 2021;175(2):176–84.

4. Lu X, Zhang L, Du H, Zhang J, Li YY, Qu J, et al. SARS-CoV-2 Infection in Children. N Engl J Med. 2020;382(17):1663–5.

5. Tsankov BK, Allaire JM, Irvine MA, Lopez AA, Sauvé LJ, Vallance BA, et al. Severe COVID-19 Infection and Pediatric Comorbidities: A Systematic Review and Meta-Analysis. Int J Infect Dis. 2021;103:246–56.

6. Gidding HF, Machalek DA, Hendry AJ, Quinn HE, Vette K, Beard FH, et al. Seroprevalence of SARS-CoV-2-specific antibodies in Sydney after the first epidemic wave of 2020. Medical Journal of Australia. 2021;214(4):179–85.

7. McRae JE, Quinn HE, Saravanos GL, Carlson SJ, Britton PN, Crawford NW, et al. Paediatric Active Enhanced Disease Surveillance (PAEDS) 2017 and 2018: Prospective hospital-based surveillance for serious paediatric conditions. Commun Dis Intell (2018). 2020;44.

8. McRae JE, Quinn HE, Saravanos GL, McMinn A, Britton PN, Wood N, et al. Paediatric Active Enhanced Disease Surveillance (PAEDS) annual report 2016: Prospective hospital-based surveillance for serious paediatric conditions. Commun Dis Intell (2018). 2019;43.

9. Wurzel D, McMinn A, Hoq M, Blyth CC, Burgner D, Tosif S, et al. Prospective characterisation of SARS-CoV-2 infections among children presenting to tertiary paediatric hospitals across Australia in 2020: a national cohort study. BMJ Open. 2021;11(11):e054510.

10. Williams P, Koirala A, Saravanos GL, Lopez LK, Glover C, Sharma K, et al. COVID-19 in New South Wales children during 2021: severity and clinical spectrum. Med J Aust. 2022;217(6):303–10.

11. Lopez L, Burgner D, Glover C, Carr J, Clark J, Boast A, et al. Lower risk of multi-system inflammatory syndrome in children (MIS-C) with the omicron variant. The Lancet Regional Health – Western Pacific. 2022;27.

12. Channon-Wells S, Vito O, McArdle AJ, Seaby EG, Patel H, Shah P, et al. Immunoglobulin, glucocorticoid, or combination therapy for multisystem inflammatory syndrome in children: a propensity-weighted cohort study. The Lancet Rheumatology. 2023;5(4):e184–e99.

13. Koirala A, Gidding HF, Vette K, Macartney K, Group tPS. The seroprevalence of SARS-CoV-2-specific antibodies in children, Australia, November 2020 – March 2021. Medical Journal of Australia. 2022;217(1):43–5.

14. Basseal JM, Bennett CM, Collignon P, Currie BJ, Durrheim DN, Leask J, et al. Key lessons from the COVID-19 public health response in Australia. The Lancet Regional Health – Western Pacific. 2023;30.

15. Australian Institute of Health and Welfare. Australia’s health 2022: data insights: Australian Institute of Health and Welfare; 2022 [updated December 23. Available from: https://www.aihw.gov.au/reports/australias-health/australias-health-2022-data-insights/about.

16. National Centre for Immunisation Research and Surveillance. Significant events in COVID-19 vaccination practice in Australia 2023 [updated November. Available from: https://ncirs.org.au/sites/default/files/2022-11/COVID-19-history-November%202022.pdf.

17. Australian COVID-19 Serosurveillance Network. Seroprevalence of SARS-CoV-2-specific antibodies among Australian blood donors: Round 4 update 2023 [updated February 8; cited 2023 June 5]. Available from: https://kirby.unsw.edu.au/sites/default/files/COVID19-Blood-Donor-Report-Round4-Nov-Dec-2022.pdf.

18. Oeser C, Whitaker H, Linley E, Borrow R, Tonge S, Brown CS, et al. Large increases in SARS-CoV-2 seropositivity in children in England: Effects of the delta wave and vaccination. J Infect. 2022;84(3):418–67.

19. Ratcliffe H, Tiley KS, Andrews N, Amirthalingam G, Vichos I, Morey E, et al. Community seroprevalence of SARS-CoV-2 in children and adolescents in England, 2019–2021. Archives of Disease in Childhood. 2023;108(2):123–30.

20. Messiah SE, DeSantis SM, Leon-Novelo LG, Talebi Y, Brito FA, Kohl HW, III, et al. Durability of SARS-CoV-2 Antibodies From Natural Infection in Children and Adolescents. Pediatrics. 2022;149(6).

21. Roche Diagnostics. Elecysis® Anti-SARS-CoV-2: F. Hoffman-La Roche Ltd; 2023 [updated June 5. Available from: https://diagnostics.roche.com/global/en/products/params/elecsys-anti-sars-cov-2.html.

22. Torres Ortiz A, Fenn Torrente F, Twigg A, Hatcher J, Saso A, Lam T, et al. The influence of time on the sensitivity of SARS-CoV-2 serological testing. Sci. 2022;12(1):10517.

23. Perez-Saez J, Zaballa M-E, Yerly S, Andrey DO, Meyer B, Eckerle I, et al. Persistence of anti-SARS-CoV-2 antibodies: immunoassay heterogeneity and implications for serosurveillance. Clinical Microbiology and Infection. 2021;27(11):1695.e7-.e12.

24. Public Health England. Evaluation of Roche Elecsys Anti-SARS-CoV-2 S serology assay for the detection of anti-SARS-CoV-2 S antibodies. 2021 March 11.

25. Riester E, Findeisen P, Hegel JK, Kabesch M, Ambrosch A, Rank CM, et al. Performance evaluation of the Roche Elecsys Anti-SARS-CoV-2 S immunoassay. J Virol Methods. 2021;297:114271.

26. Wang W, Rothschild D, Goel S, Gelman A. Forecasting elections with non-representative polls. International Journal of Forecasting. 2015;31(3):980–91.

27. Gelman A, Carpenter B. Bayesian Analysis of Tests with Unknown Specificity and Sensitivity. J R Stat Soc Ser C Appl Stat. 2020;69(5):1269–83.

28. Australian Government Department of Health. National Notifiable Diseases Surveillance System (NNDSS) datasets. 2023.

29. Australian Bureau of Statistics. Quarterly Population Estimates (ERP), by State/Territory, Sex and Age, 2022-Q3. 2023.

30. Wilde H, Tomlinson C, Mateen BA, Selby D, Kanthimathinathan HK, Ramnarayan P, et al. Hospital admissions linked to SARS-CoV-2 infection in children and adolescents: cohort study of 3.2 million first ascertained infections in England. BMJ. 2023;382:e073639.

31. Bobrovitz N, Ware H, Ma X, Li Z, Hosseini R, Cao C, et al. Protective effectiveness of previous SARS-CoV-2 infection and hybrid immunity against the omicron variant and severe disease: a systematic review and meta-regression. Lancet Infect Dis. 2023;23(5):556–67.

32. Rotulo GA, Palma P. Understanding COVID-19 in children: immune determinants and post-infection conditions. Pediatr Res. 2023;94(2):434–42.

33. Cambou MC, Liu CM, Mok T, Fajardo-Martinez V, Paiola SG, Ibarrondo FJ, et al. Longitudinal Evaluation of Antibody Persistence in Mother-Infant Dyads After Severe Acute Respiratory Syndrome Coronavirus 2 Infection in Pregnancy. The Journal of Infectious Diseases. 2022;227(2):236–45.

34. Jorgensen SCJ, Hernandez A, Fell DB, Austin PC, D’Souza R, Guttmann A, et al. Maternal mRNA covid-19 vaccination during pregnancy and delta or omicron infection or hospital admission in infants: test negative design study. BMJ. 2023;380:e074035.

35. Steinhoff MC, Omer SB, Roy E, Arifeen SE, Raqib R, Altaye M, et al. Influenza Immunization in Pregnancy — Antibody Responses in Mothers and Infants. N Engl J Med. 2010;362(17):1644–6.

36. Nunes MC, Cutland CL, Jones S, Hugo A, Madimabe R, Simões EAF, et al. Duration of Infant Protection Against Influenza Illness Conferred by Maternal Immunization: Secondary Analysis of a Randomized Clinical Trial. Jama, Pediatr. 2016;170(9):840–7.

37. Otto M, Britton PN, Serpa Neto A, Erickson S, Festa M, Crawford NW, et al. COVID-19 related ICU admissions in paediatric and young adult patients in Australia: a national case series 2020-2022. Lancet Reg Health West Pac. 2023;36:100763.

38. Hobbs CV, Woodworth K, Young CC, Jackson AM, Newhams MM, Dapul H, et al. Frequency, Characteristics and Complications of COVID-19 in Hospitalized Infants. The Pediatric Infectious Disease Journal. 2022;41(3):e81–e6.

39. Halasa NB, Olson SM, Staat MA, Newhams MM, Price AM, Pannaraj PS, et al. Maternal Vaccination and Risk of Hospitalization for Covid-19 among Infants. N Engl J Med. 2022;387(2):109–19.

40. Van Elslande J, Oyaert M, Lorent N, Vande Weygaerde Y, Van Pottelbergh G, Godderis L, et al. Lower persistence of anti-nucleocapsid compared to anti-spike antibodies up to one year after SARS-CoV-2 infection. Diagn Microbiol Infect Dis. 2022;103(1):115659.

41. Navaratnam AMD, Shrotri M, Nguyen V, Braithwaite I, Beale S, Byrne TE, et al. Nucleocapsid and spike antibody responses following virologically confirmed SARS-CoV-2 infection: an observational analysis in the Virus Watch community cohort. Int J Infect Dis. 2022;123:104–11.

42. Ireland G, Jeffery-Smith A, Zambon M, Hoschler K, Harris R, Poh J, et al. Antibody persistence and neutralising activity in primary school students and staff: Prospective active surveillance, June to December 2020, England. eClinicalMedicine. 2021;41.

43. Nakagama Y, Komase Y, Kaku N, Nitahara Y, Tshibangu-Kabamba E, Tominaga T, et al. Detecting Waning Serological Response with Commercial Immunoassays: 18-Month Longitudinal Follow-up of Anti-SARS-CoV-2 Nucleocapsid Antibodies. Microbiol Spectr. 2022;10(4):e0098622.

44. Australian Bureau of Statistics. Australian population grid 2022 2022 [updated April 20, 2023. Available from: https://www.abs.gov.au/statistics/people/population/regional-population/latest-release.

45. Australian Institute of Health and Welfare. Data tables: ABDS 2022 National estimates for Australia 2022 [updated December 14, 2022. Available from: https://www.aihw.gov.au/reports-statistics/health-conditions-disability-deaths/burden-of-disease/overview.

